# Morphological and Functional Alterations in Type 2 Diabetes Pancreata assessed with MRI-based metrics and [^18^F]FP-(+)-DTBZ PET

**DOI:** 10.1101/2025.10.13.25337899

**Authors:** Seyed Faraz Nejati, Faranak Ebrahimian Sadabad, Rui Ren, Yuan Huang, Jason Bini

**Author notes:** Corresponding Author: Jason Bini, PhD Assistant Professor PET Center Yale Biomedical Imaging Institute Department of Radiology and Biomedical Imaging Yale University 801 Howard Avenue New Haven, CT 06520.

## Abstract

**Objective:** To determine if combining PET-derived beta-cell mass (BCM) estimates with MRI-based morphology metrics improves the prediction of beta-cell functional mass in type 2 diabetes (T2D).

**Methods:** We performed a retrospective analysis of 40 participants; 19 T2D, 16 healthy obese volunteers (HOV), 5 prediabetes, who underwent [^18^F]FP-(+)-DTBZ PET to quantify vesicular monoamine transporter type 2 (VMAT2) density (SUVR-1), T1-weighted MRI for 3D morphology metric analysis, and an arginine stimulus test to measure acute (AIRarg) and maximum (AIRargMAX) insulin responses. Lasso regression models identified the optimal combination of PET, MRI, and clinical variables to predict beta-cell function for the whole pancreas and its subregions.

**Results:** Compared to HOV, individuals with T2D exhibited significantly reduced AIRarg and AIRargMAX. Only pancreas body volume was significantly smaller in the T2D cohort. For the whole pancreas, a model including PET-derived SUVR-1 and a subset of clinical covariates best predicted acute beta-cell function (AIRarg). However, predicting maximum functional reserve (AIRargMAX) required the addition of MRI-based morphology metrics in combination with SUVR-1 and a subset of clinical covariates.

**Conclusion:** We combined PET imaging of BCM and MRI morphology metrics with a robust machine learning-based variable selection method to extract useful PET- and MRI-based metrics for predicting functional and not-fully functional BCM. This synergistic approach offers a novel combination of biomarkers for staging disease and evaluating therapeutic interventions.

## Introduction

Type 2 diabetes (T2D) is characterized by chronic insulin resistance, increased β-cell workload to failure and eventual decline in β-cell function and mass in the pancreas[1]. Several imaging studies have demonstrated pancreatic volume loss ranging from 13-33%[2–5]. Along with pancreatic volume loss, autopsy studies have demonstrated loss of 40-65% of beta cell mass (BCM) in individuals with T2D[6,7]. Only 1-3% of pancreas volume consists of islet mass; therefore, understanding the mechanisms of pancreatic volume loss, in both the endocrine and exocrine pancreas in T2D is important.

Several biomarkers are currently used to assess endocrine and exocrine function in the pancreas[8–11]. Endocrine pancreas function can be assessed using peripheral blood measurements such as Hemoglobin A1c (HbA1c), insulin, C-peptide, and proinsulin, as well as the ratio of proinsulin to C-peptide (PI:C ratio)[8–10]. More rigorous tests of functional BCM can be performed, such as the arginine stimulus test (AST)[12,13]. Intravenous arginine is an ideal β- and α-cell agonist that allows simultaneous examination of insulin, C-peptide, and glucagon responses[12–14]. Exocrine pancreas secretory enzymes such as amylase, lipase and trypsinogen have been proposed as serological biomarkers with relationships to pancreatic volume loss[11]. Although recent studies have demonstrated the clinical utility of serum biomarkers, such as PI:C ratio, to assess treatment response[10], further understanding of the relationship between serum biomarkers and both endocrine and exocrine pancreas structure and function is necessary.

To understand the fate of beta cells during diabetes, measurement of BCM *in vivo* is largely done using positron emission tomography (PET) imaging with radioligands that bind primarily to receptor targets on beta cells. There are several targets that are currently being pursued including vesicular monoamine transporter type 2 (VMAT2), dopamine receptors, and GLP-1 receptors[3,15–18]. VMAT2, primarily expressed within beta cells, is a transmembrane protein responsible for sequestering insulin and dopamine into insulin-secretory granules to regulate insulin secretion[19]. [^18^F]fluoropropyl-dihydrotetrabenazine ([^18^F]FP-(+)-DTBZ), is a radioligand that binds to VMAT2 and has been shown to highly correlate with BCM[3,20–26]. Initial human studies using [^18^F]FP-(+)-DTBZ to measure BCM were performed in patients with type 1 diabetes (T1D) [21,23]; however, recently this method was extended to patients with type 2 diabetes (T2D)[3]. In the T2D study, correlations of VMAT2 density, a biomarker of BCM, correlated with years of T2D diagnosis, glycemic control and beta cell functional measures suggesting that PET was able to quantify changes in BCM[3]. Magnetic resonance imaging (MRI) in the same subjects revealed a pancreatic volume decrease of ∼13% in T2D, compared to healthy obese volunteers (HOV), suggesting loss of both endocrine BCM and exocrine volume.

Cross-sectional and longitudinal studies in patients with T1D or T2D have examined the role of pancreas volume or pancreatic volume index (PVI), volume normalized to body weight [27–30]. More recently, the identification of pancreas morphology metrics; for example, surface area, long and short axis lengths, and ratio of longest to shortest axes, have revealed more complex changes in pancreas structure beyond pancreas volume and PVI in longitudinal studies of patients with T1D[29,30]. Combining PVI and pancreas morphology metrics classification of individuals with T1D versus healthy controls improved compared to using only PVI[29,30]. Similar MRI-based morphology metrics have been proposed in T2D[2,4,31].

To our knowledge no one has combined PET BCM measurements with pancreas morphology metrics, beyond pancreas volume or PVI, as was done previously[3]. In this retrospective study, we re-examined PET and MRI data with the addition of new pancreas morphology metrics to reveal important endocrine and exocrine pancreas features that may predict BCM and function in T2D. This was done by reporting pancreas MRI morphology metrics in healthy obese volunteers (HOV) and patients with T2D and combining MRI and PET metrics in a logistic regression that predicted functional BCM. Exploratory group difference analyses and linear correlations also revealed that both PET and MRI morphology metrics can predict aspects of beta cell mass and function in T2D.

## Materials and Methods

This is a retrospective analysis of the previously published [^18^F]FP-(+)-DTBZ PET imaging study[3]. The study was approved by the Yale University Human Investigation Committee and the Yale-New Haven Hospital Radiation Safety Committee and in accordance with federal guidelines and regulations of the USA for the protection of human research subjects contained in Title 45 Part 46 of the Code of Federal Regulations (45 CFR 46). All participants signed a written informed consent.

Briefly, the original study included 40 participants: 16 HOV, five individuals with prediabetes, and 19 individuals with T2D. All subjects underwent pancreas [^18^F]FP-(+)-DTBZ PET, pancreas MRI acquisition and an arginine stimulus test (AST). AST provided two outcome measures, acute insulin response to arginine (AIRarg, serum C-peptide from 0-5 min) and maximum insulin response to arginine (AIRargMAX, serum C-peptide from 55-65 min)[12,13]. Further details regarding PET and MR acquisition, image reconstruction, AST protocol, and quantitative PET analyses can be found in the previous manuscript[3].

For the current goal of examining the effectiveness of PET and MRI imaging metrics to predict BCM, we included three outcomes: AIRarg (functional beta cell mass), AIRargMAX (functional and not fully functional beta cell mass) and the ratio of acute to maximum insulin response to arginine (acute:MAX), reflective of the ratio of functional to functional and not fully functional beta cell mass. Additional clinical measures included: age, gender, weight, BMI, hemoglobin A1c (HbA1c), years of diabetes and diagnosis (*e.g.*, HOV, prediabetes, or T2D).

PET surrogate outcome measures of BCM include the non-displaceable binding potential (*BP*_ND_)[32] and standardized uptake value ratio (SUVR-1), both using the spleen as a reference region to account for non-specific radiotracer uptake[3]. *BP*_ND_ *x* pancreas volume and SUVR-1 *x* pancreas volume can also be calculated to account for organ volume loss and is reflective of *aggregate* pancreas BCM.

For the current study, we performed manual pancreas segmentation of the whole pancreas on a the T1-weighted abdominal MRI[3]. The researcher performing the pancreas segmentation was blinded diagnosis of each participant. Following previous methods[29], whole pancreas regions-of-interest (ROIs) were drawn in the axial plane using Medical Image Processing, Analysis and Visualization (MIPAV) software Center for Information Technology, National Institutes of Health, version 11.0.7-2023-06-22, https://mipav.cit.nih.gov). ROIs were also subdivided into pancreas head, body and tail. For each subject, the stack of axial pancreas whole, head, body and tail ROIs were converted to their respective volumes of interest and subsequently to a 3-dimensional binary mask of the pancreas in MIPAV. This 3D binary mask was then used as input into ‘regionprops3’ (MATLAB, version R2023a, The MathWorks, https://www.mathworks.com/products/matlab) to calculate MRI morphology metrics of the 3D pancreas mask. MRI morphology metric outputs from ‘regionprops3’ include: 1) ‘BoundingBox’, the smallest cuboid containing the pancreas with lengths ‘BoundingBox1’, BoundingBox2’, BoundingBox3’ and ‘BoundingBoxVolume’, 2) ‘Centroid’, coordinates of the center of mass of the pancreas (centroid1, centroid2 and centroid3), 3) EquivDiameter, diameter of a sphere with the same volume as the pancreas, 4) Extent, ratio of voxels in the pancreas to voxels in the total bounding box, 5) PrincipalAxisLength, length in voxels of the major axes of the ellipsoid that have the same normalized second central moments as the pancreas (PrincipalAxisLength1, PrincipalAxisLength2, PrincipalAxisLength3), 6) ConvexVolume, number of voxels in the smallest convex polygon that contains the pancreas, 7) Solidity, proportion of voxels in the convex volume that are also in the pancreas, 8) pancreas surface area, and 9) pancreas volume. All metrics were calculated separately for the whole pancreas, and pancreas head, body and tail.

### Statistical Analysis

One-way ANOVA for each functional BCM outcome (AIRarg, AIRargMAX, and acute:MAX) was performed and when appropriate group differences are compared with an unpaired t test with Welch’s correction. We also aimed to investigate the relationships between functional BCM outcomes (AIRarg, AIRargMAX, and acute:MAX) and various predictors (PET variables, MRI morphology metrics, and clinical covariates (*e.g.*, age, BMI) (**Figure 1**). For each pancreas ROI delineation (whole, head, body, or tail), we constructed 3 models, each incorporating one PET variable, all MRI morphology metrics and all clinical covariates for a specific functional BCM outcome. Previously[3], SUVR-1 was the primary outcome variable; therefore, we used that as our primary PET outcome measure (**Figure 1**, **yellow circles**).

**Figure 1.**
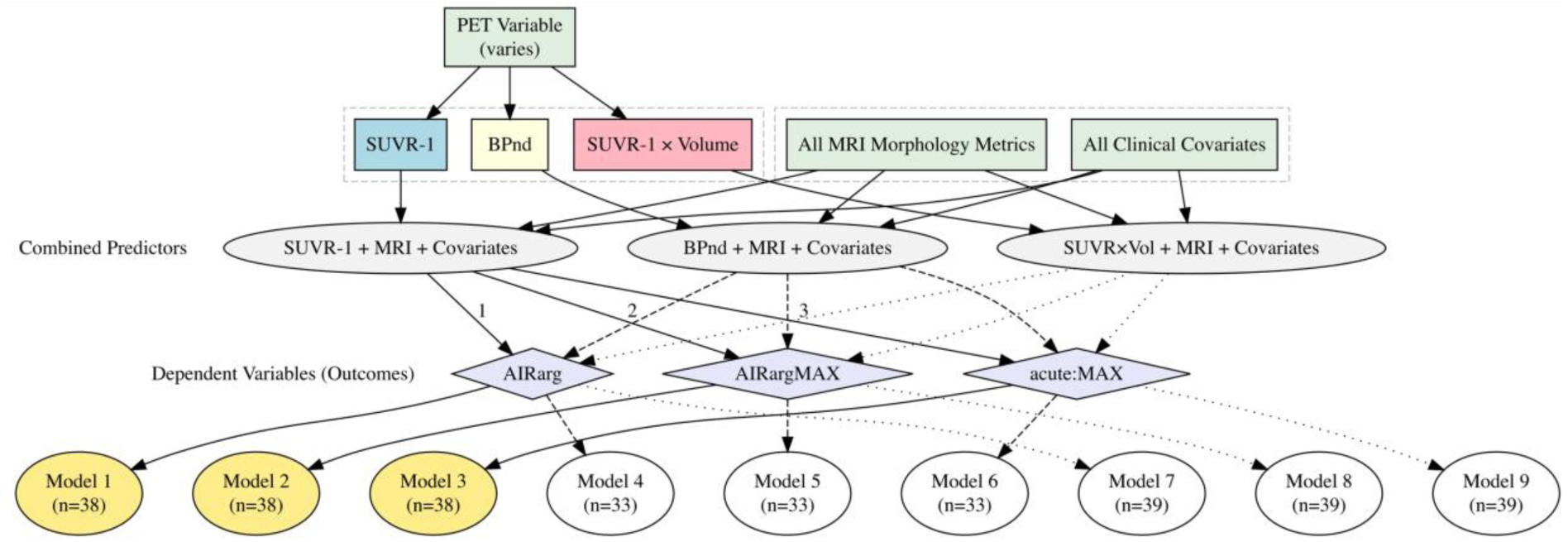
Predictors and Outcome Composition in Full Linear Regression Models. SUVR-1 (blue box) was the primary PET outcome metric used to predict outcomes (purple diamonds). Each model contained combined predictors (gray ovals) consisting of a PET variable, all MRI morphology metrics and all clinical covariates (green rectangles). Primary outcome models used SUVR-1, all MRI morphology metrics and all clinical covariates to predict functional BCM outcomes (Models 1-3, yellow circles). Exploratory analyses used *BP*ND or SUVR-1 × Volume with all MRI morphology metrics and all clinical covariates (Models 4-9).

Additional models were examined as exploratory analyses, using alternate PET outcomes (*BP*_ND_) or pancreas aggregate binding measures (SUVR-1 × Volume) (**Figure 1**, **white circles**).

Differences in sample sizes in each model arose from removing cases with missing data in various combinations of these variables.

### Predictive Modeling with Variable Selection

The primary goal was to determine whether a subset of variables could achieve prediction accuracy comparable to or better than a model using all predictors. To identify such a subset, we applied Lasso regression with bootstrap-based variable-selection procedure to achieve more stable performance. Lasso regression is a commonly used approach in high-dimensional settings that identifies variables with nonzero coefficients, indicating potential relevance to the outcome[33]. Given the relatively small sample size and the large number of predictors, we implemented a bootstrap strategy to improve selection stability. Specifically, we generated B=500 bootstrap samples and recorded the predictors selected in each iteration. For each variable, we calculated its selection frequency as f=C/B, where C represents the number of times the variable was selected across all resamples. Variables with f > 0.5 were retained for inclusion in the final reduced model.[34]

To evaluate prediction performance, we implemented Leave-One-Out Cross Validation (LOOCV). This involved training the model on all but one observation and calculating the squared prediction error, (*y* − *ŷ*)^2^, for the omitted observation. The average of these errors yielded the mean squared error (MSE). We then compared the MSE of the full model (using all predictors) with that of a reduced model (using only selected variables with f > 0.5).

## Results

One-way ANOVA demonstrated significant differences in the group means for each of the functional BCM outcomes AIRarg (p=0.04), AIRargMAX (<0.0001), and acute:MAX (p=0.002); therefore, we examined group differences of the respective outcome measures between HOV, prediabetic and T2D (**Figure 2**).

**Figure 2.**
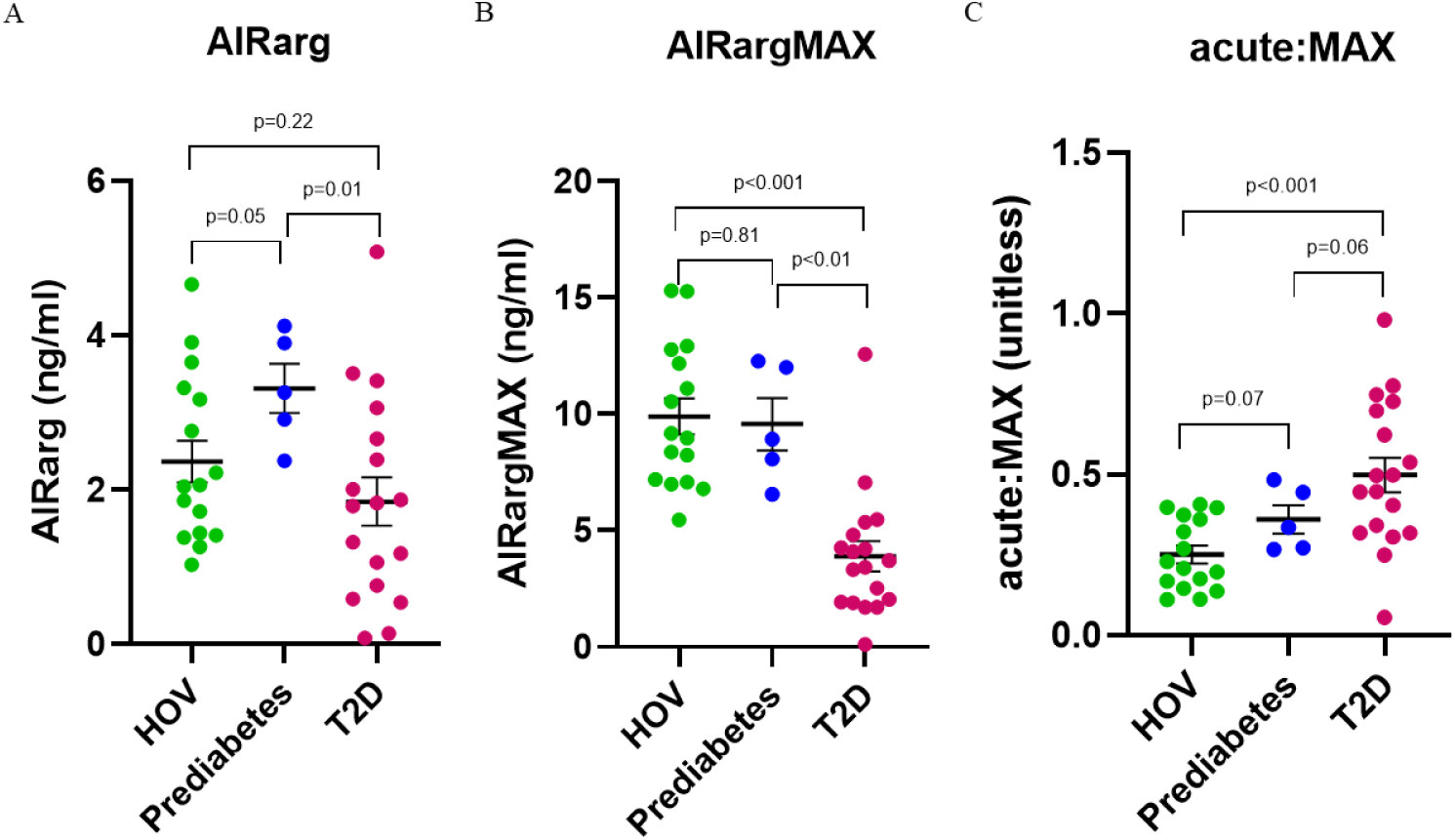
Group comparisons of functional beta cell mass outcomes **A)** Acute insulin response to arginine stimulus (AIRarg) **B)** Maximum insulin response to arginine (AIRargMAX) **C)** ratio of acute to maximum response to arginine (acute:MAX). All data presented as mean ± SEM.

AIRarg, is higher in those who have prediabetes (mean ± SEM: 3.3 ± 0.3 ng/ml) than the HOV group (2.4 ± 0.3 ng/ml) but is significantly lower than both groups in the patients with T2D (1.9 ± 0.3 ng/ml, p=0.01) (**Figure 2A**). AIRargMAX, is similar in the HOV (9.9 ± 0.8 ng/ml) and prediabetes (9.6 ± 1.1 ng/ml) groups, while in patients with T2D it is significantly lower (3.9 ± 0.7 ng/ml, p<0.01) (**Figure 2B**). For acute:MAX, the values are in rank order, from low to high, between groups HOV (0.25 ± 0.03 unitless) < prediabetes (0.36 ± 0.04 unitless) < T2D (0.50 ± 0.05 unitless); however, only the difference between HOV and T2D is significant (p<0.001) (**Figure 2C**).

Representative whole pancreas axial slices of HOV (**Figure 3A**) and T2D (**Figure 3B**) as visualized by MRI. Pancreas ROIs in both HOV (**Figure 3C**) and T2D (**Figure 3D**) subdivided into pancreas head (blue outline), body (red outline), and tail (green outline) ROIs.

**Figure 3.**
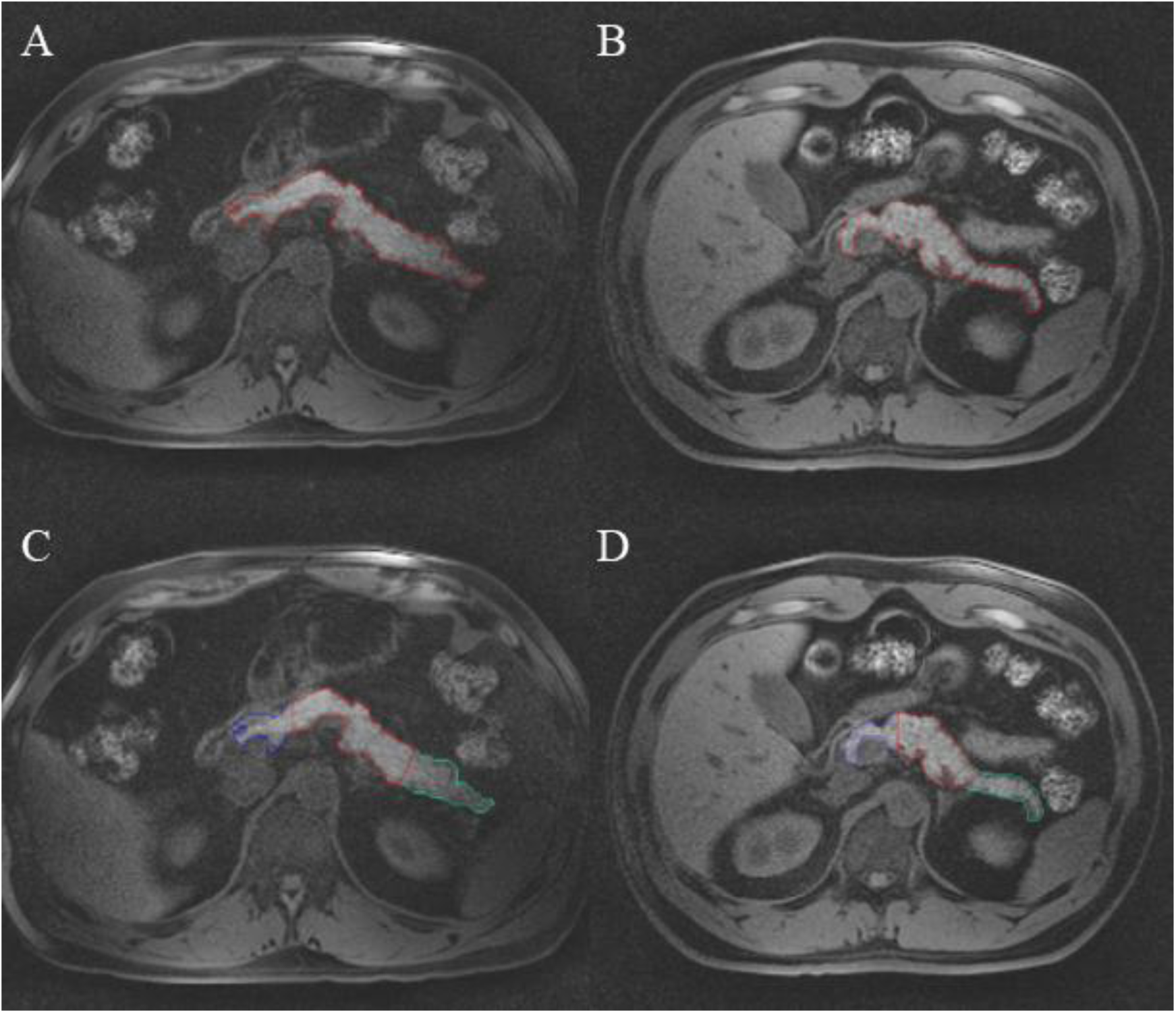
Representative axial pancreas MRI images of whole pancreas region-of-interest in a **A)** healthy obese volunteer and **B)** individual with type 2 diabetes. Subdivision of pancreas regions-of-interest for head (blue), body (red), and tail (green) in **C)** a healthy obese volunteer and **D)** an individual with T2D.

A representative stack of axial pancreas whole, head, body and tail ROIs from a HOV and individual with T2D converted to their respective 3D binary masks (**Figure 4**).

**Figure 4.**
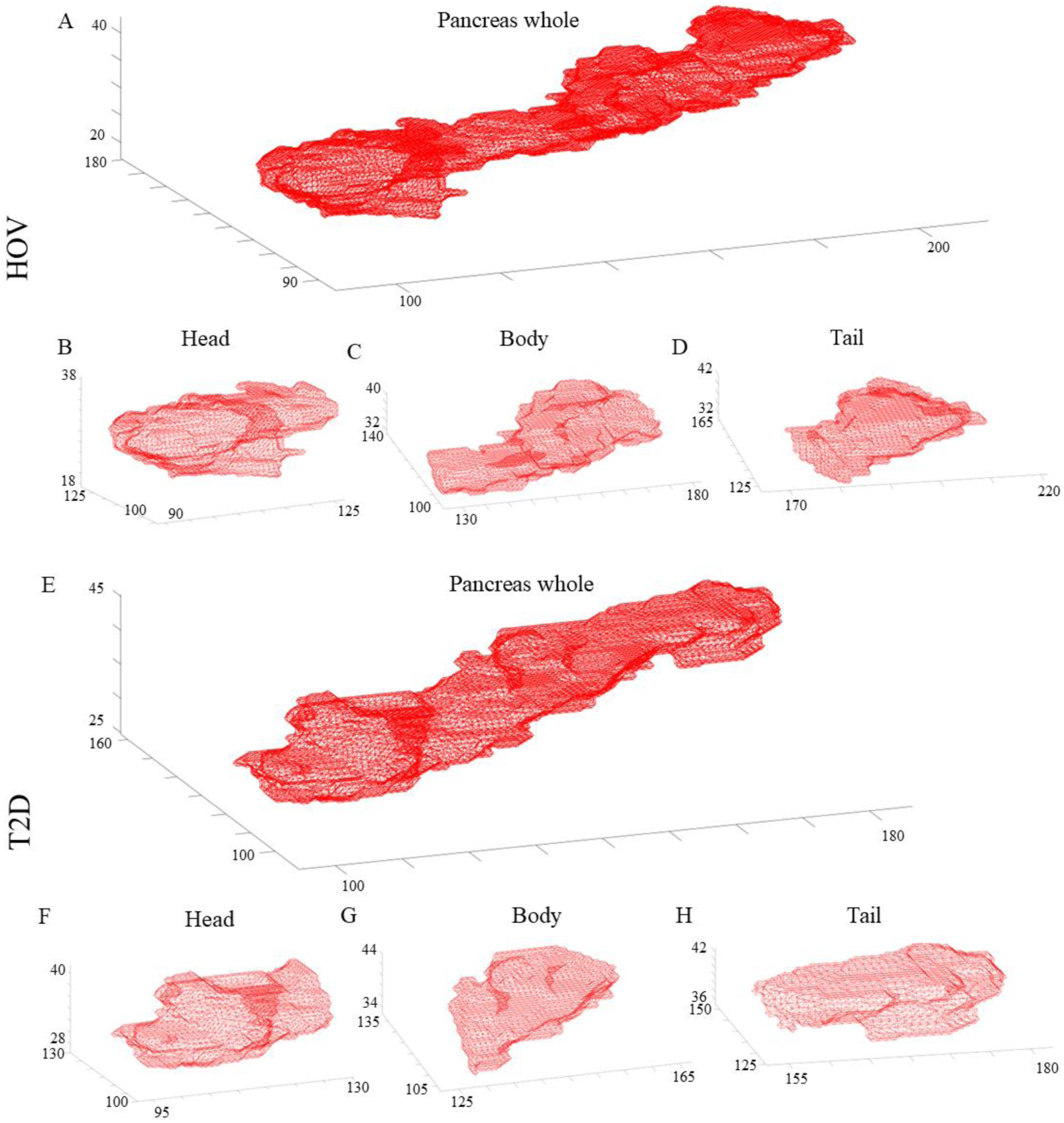
Representative 3D pancreas masks in a healthy obese volunteer; **A)** whole pancreas and pancreas **B)** head **C)** body and **D)** tail and an individual with type 2 diabetes; **E)** whole pancreas and pancreas **F)** head **G)** body and **H)** tail. All axes are voxel numbers. For example, in **A)** the healthy obese volunteer the whole pancreas would be roughly be bounded by a rectangle of size 100 x 90 x 20 voxels.

MRI morphology metric outcomes in a HOV from ‘regionprops’ are presented to demonstrate their relationship to pancreas morphology (**Figure 5**).

**Figure 5.**
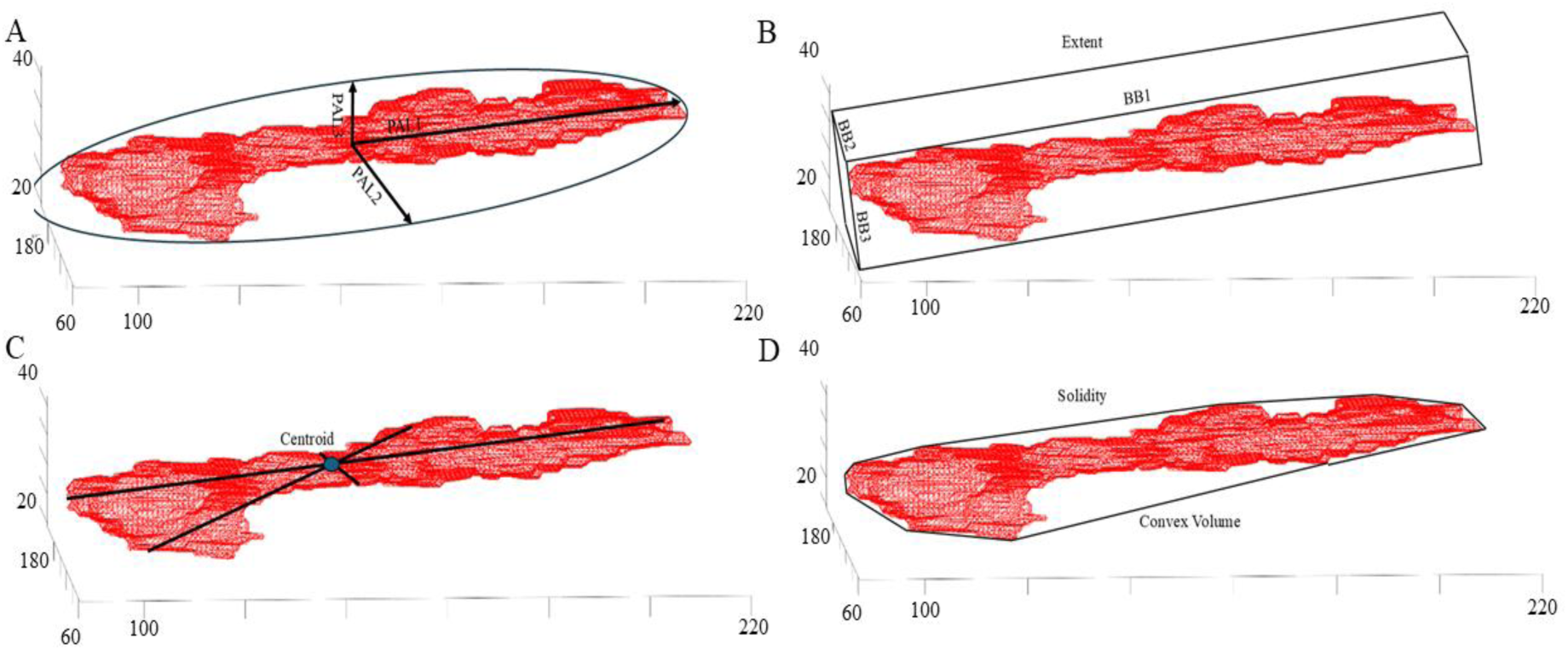
MRI morphology metric outputs from ‘regionprops3’ including: **A)** Principal Axis Lengths (PAL) of the smallest ellipsoid encapsulating the pancreas **B)** Smallest Bounding Box (BB) surrounding the pancreas and Extent, the ratio of voxels in the pancreas to voxels in the Bounding Box **C**) Coordinates (x,y,z) of the center of mass of the pancreas (Centroid) and **D)** Smallest Convex Volume surrounding the pancreas and Solidity, the proportion of voxels in the convex volume that are also in the pancreas region.

No group differences were seen in any pancreas ROI using SUVR-1, as previously reported (**Supplementary Figure 1**)[3]. The pancreas body volume was the only ROI that was significantly different between HOV (mean ± SEM; 39.1 ± 3.4 ml) and individuals with T2D (28.8 ± 3.0 ml, p=0.01) (**Supplementary Figure 1**).

Regression models were analyzed to investigate the relationship between functional beta cell mass outcomes: AIRarg, AIRargMAX, and acuteMAX and predictive variables, including the primary PET metric (SUVR-1), MRI morphology metrics, and clinical covariates for whole pancreas and subregions of the pancreas including head, body and tail (**Table 1**). For all linear models and regions, the reduced linear model had lower MSE (**Figure 6**), indicating that the functional BCM outcomes could be better predicted by a specific subset of PET, MRI and/or clinical covariates.

**Figure 6.**
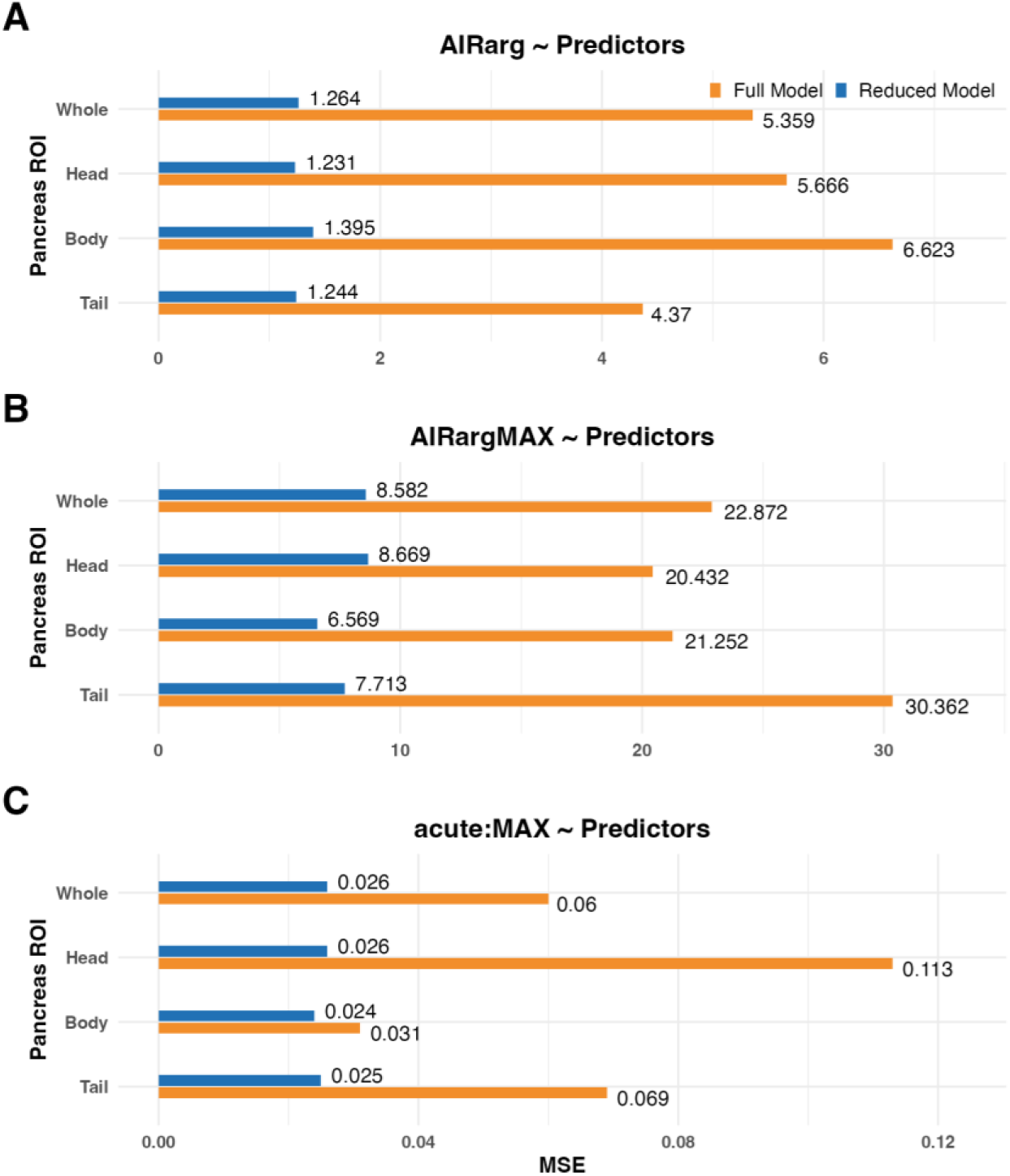
Comparison of mean square error of full (orange bars) and reduced (blue bars) models used to predict functional β-cell mass outcomes: **A)** AIRarg **B)** AIRargMAX and **C)** acute:MAX across pancreas and subregions.

**Table 1.** Reduced models for predicting primary functional beta cell mass outcomes (AIRarg, AIRargMAX and acute:MAX) with the primary PET outcome measure (SUVR-1), MRI morphology metrics and clinical covariates.

| PET outcome measure | Pancreas ROI | Linear Models to Predict Acute Insulin Response to Arginine (AIRarg) |
| --- | --- | --- |
| SUVr-1 | Whole | AIRarg ~ SUVr-1 + age + gender + years.of.diabetes + diagnosispreDM |
|  | Head | AIRarg ~ SUVr-1 + age + gender + years.of.diabetes + diagnosispreDM |
|  | Body | AIRarg ~ SUVr-1 + PrincipalAxisLength3 + Solidity + age + years.of.diabetes + diagnosispreDM |
|  | Tail | AIRarg ~ SUVr-1 + BoundingBox1 + age + gender + years.of.diabetes + diagnosispreDM |
|  |  | Linear Models to Predict maximum insulin response to arginine (AIRargMAX) |
| SUVr-1 | Whole | AIRargMAX ~ SUVr-1 + Centroid1 + Centroid2 + Centroid3 + PrincipalAxisLength3 + gender + HbA1c + years.of.diabetes + diagnosisT2D |
|  | Head | AIRargMAX ~ SUVr-1 + Centroid2 + Centroid3 + PrincipalAxisLength2 + PrincipalAxisLength3 + age + gender + HbA1c + years.of.diabetes + diagnosisT2D |
|  | Body | AIRargMAX ~ SUVr-1 + Centroid1 + Centroid2 + Centroid3 + PrincipalAxisLength2 + PrincipalAxisLength3 + age + gender + HbA1c + years.of.diabetes + diagnosisT2D |
|  | Tail | AIRargMAX ~ SUVr-1 + Centroid1 + Centroid2 + Centroid3 + gender + HbA1c + years.of.diabetes + diagnosisT2D |
|  |  | Linear Models to Predict ratio of acute to maximum insulin response to arginine (acute:MAX) |
| SUVr-1 | Whole | acute:MAX ~ SUVr-1 + Centroid1 + Centroid3 + BoundingBox1 + BoundingBox3 + PrincipalAxisLength1 + PrincipalAxisLength2 + PrincipalAxisLength3 + age + gender + HbA1c + years.of.diabetes + diagnosisT2D + diagnosispreDM |
|  | Head | acute:MAX ~ SUVr-1 + PrincipalAxisLength1 + PrincipalAxisLength2 + PrincipalAxisLength3 + age + HbA1c + years.of.diabetes + diagnosisT2D + diagnosispreDM |
|  | Body | acute:MAX ~ SUVr-1 + Centroid1 + Centroid2 + Centroid3 + BoundingBox1 + BoundingBox3 + Extent + PrincipalAxisLength2 + Solidity + age + gender + weight + HbA1c + years.of.diabetes + diagnosisT2D + diagnosispreDM |
|  | Tail | acute:MAX ~ Centroid1 + Centroid2 + PrincipalAxisLength2 + age + HbA1c + years.of.diabetes + diagnosisT2D + diagnosispreDM |

In the reduced models for predicting AIRarg, SUVR-1 was included for the whole pancreas and every subregion (**Table 1**). No MRI morphology metrics were included in the reduced models for whole pancreas or head. In the pancreas body, principal axis length 3 and solidity were included, while in the tail, only bounding box 1 was included in the reduced model.

For reduced models predicting AIRargMAX, SUVR-1 was again included for whole pancreas and each subregion. For MRI morphology metrics, whole pancreas was comprised of centroid 1-3 and principal axis length 3. The head and body included different combinations of centroid 1-3 and principal axis length 2 and 3. For the tail, only centroid 1-3 were included.

In the final model, to predict acute:MAX, SUVR-1 was included in all models, except for the reduced model in the tail. The MRI morphology metrics included contain several variations, dependent on pancreas region, of the following parameters: centroid 1-3, bounding box 1 and 3, principal axes 1-3, extent and solidity.

Exploratory reduced models, using alternate PET outcomes (*BP*_ND_) or pancreas volume aggregate binding measures (SUVR-1 *x* Volume or *BP*_ND_ *x* Volume) showed similar patterns of reduced MSE and combinations of PET and MRI morphology metrics (**Supplementary Table 1 and 2**).

Exploratory analyses examining group differences (mean ± SEM) of pancreas MRI morphology metrics revealed significant differences in whole pancreas centroid 1 (HOV: 140.7 ± 1.2, T2D: 136.9 ± 1.3; p=0.04), pancreas body principal axis length 3 (HOV: 8.0 ± 0.5, T2D: 6.7 ± 0.3; p=0.01), pancreas body EquivDiameter (HOV: 18.8 ± 0.7, T2D: 16.8 ± 0.6; p=0.01), pancreas body bounding box 1 (HOV: 40.5 ± 2.2, T2D: 35 ± 1.9; p=0.04), pancreas body convex volume (HOV: 65.6 ± 5.6, T2D: 48.1 ± 5.7; p=0.01), and pancreas body surface area (HOV: 25.5 ± 1.8, T2D: 19.6 ± 1.6; p=0.01) (**Figure 7**).

**Figure 7.**
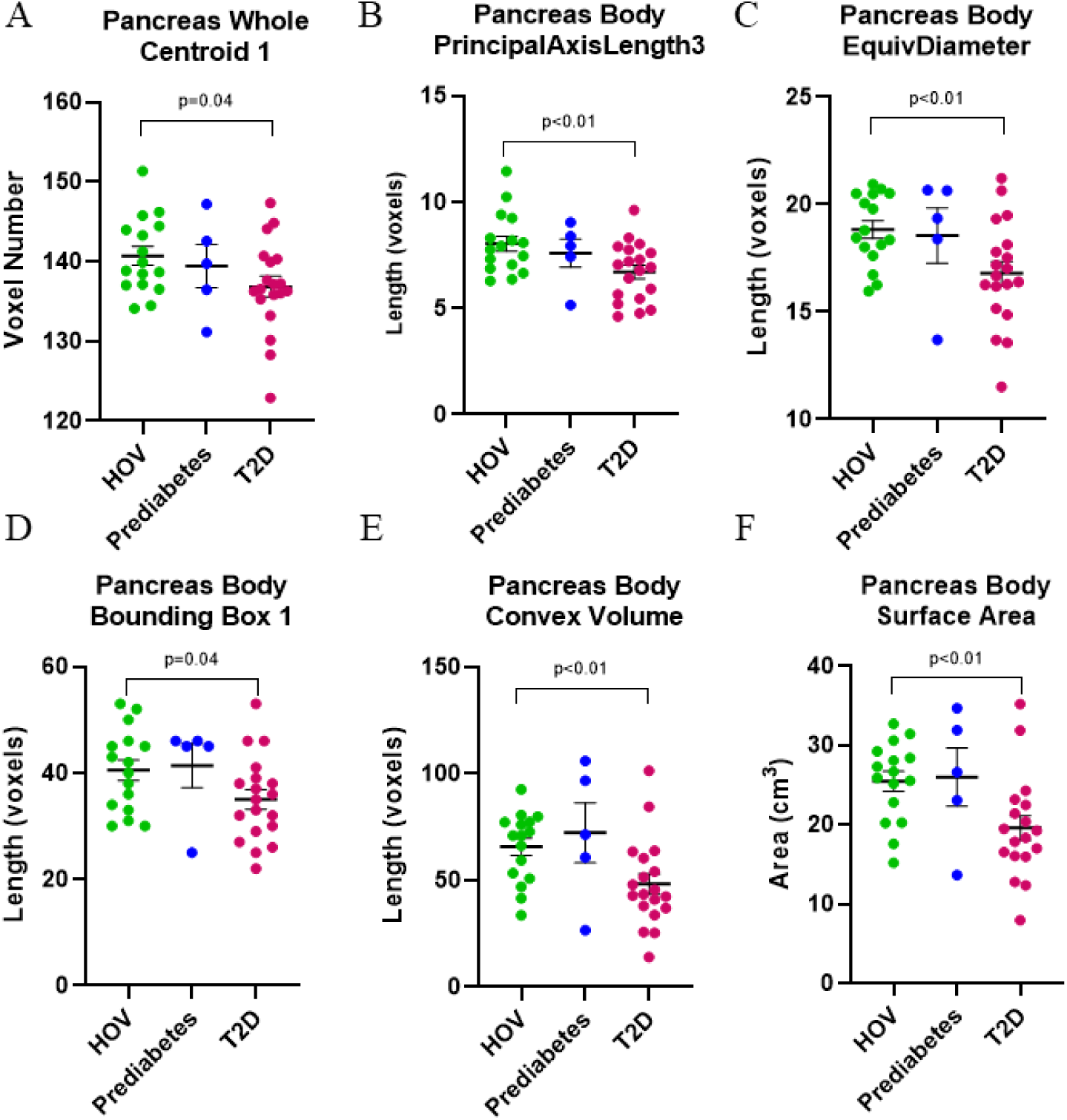
Exploratory group comparisons of pancreas MRI morphology metrics between healthy obese volunteers and individuals with T2D in whole pancreas for **A)** Centroid 1, and in pancreas body for **B)** Principal Axis Length 3 **C)** EquivDiameter **D)** Bounding Box 1 **E)** Convex Volume and **F)** Surface Area. All data presented as mean ± SEM.

Exploratory correlations between AIRarg, AIRargMAX, acute:MAX and single PET and MRI morphology metrics are displayed to compare differences and similarities of PET and MRI morphology metrics to functional BCM outcome measures (**Figure 8**).

**Figure 8.**
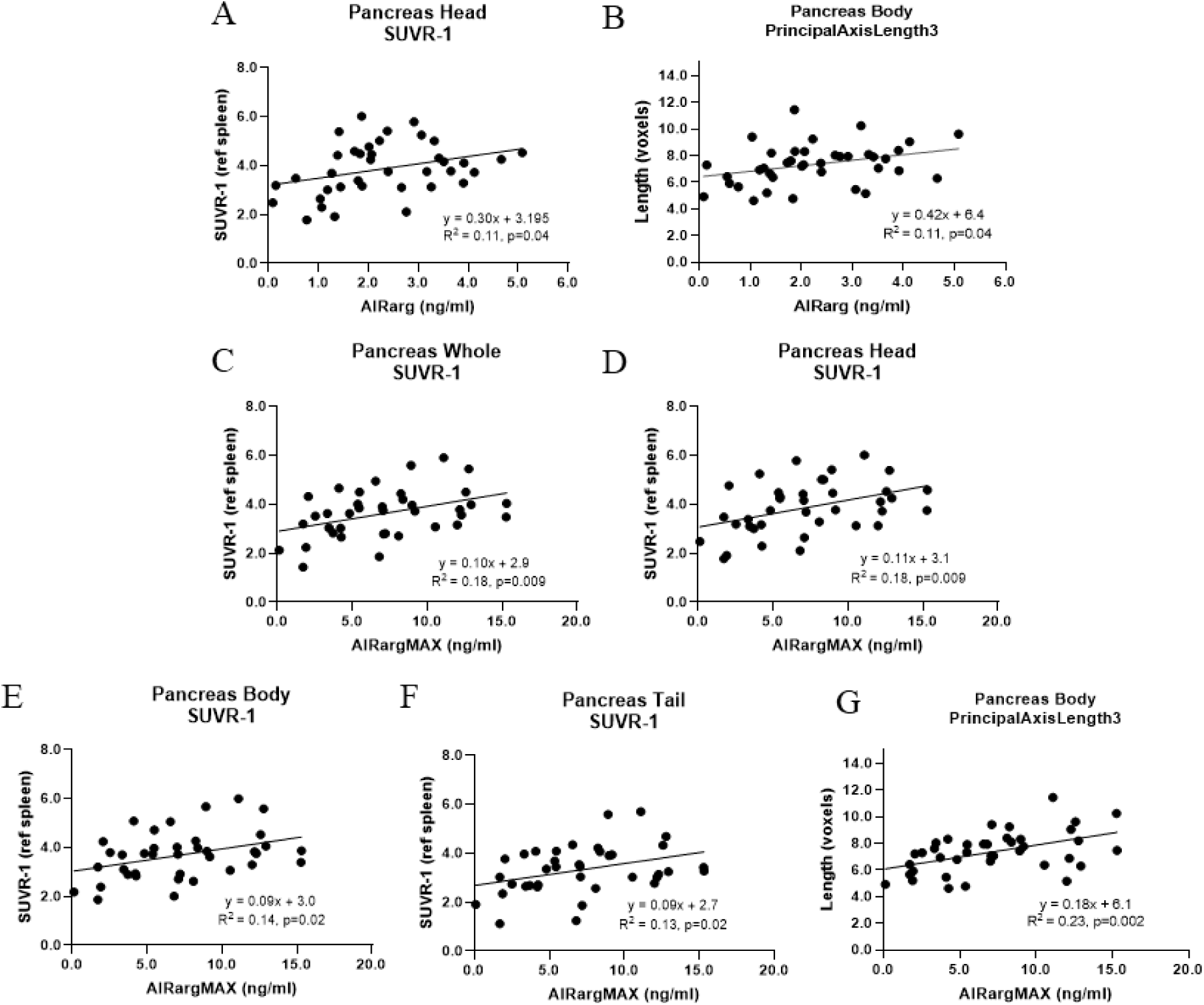
Exploratory correlations between functional beta cell mass outcome and imaging metrics. Correlations of AIRarg with A) pancreas head SUVR-1 and B) pancreas body principal axis length 3. Correlations of AIRargMAX with SUVR-1 in all pancreas regions C) whole D) head E) body and F) tail, as well as G) pancreas body principal axis length 3.

AIRarg correlated with both pancreas head SUVR-1 (R^2^=0.11, p=0.04) and pancreas body PAL3 (R^2^=0.11, p=0.04). As previously reported[3], AIRargMAX was significantly correlated with SUVR-1 in all pancreas regions; whole (R^2^=0.18, p=0.009), head (R^2^=0.18, p=0.009), body (R^2^=0.14, p=0.02) and tail (R^2^=0.13, p=0.02). However, AIRargMAX was also significantly correlated with the MRI morphology metric pancreas body principal axis length 3 (R^2^=0.23, p=0.002). No standalone imaging metrics were correlated with acute:MAX.

## Discussion

We performed a retrospective analysis of PET and MRI pancreas imaging data with new analyses of MRI morphology metrics to determine which combination of imaging-based metrics best predicts beta cell mass and function in patients with T2D.

Functional beta cell mass assessments showed significant differences between HOV and patients with T2D for all three metrics: acute (AIRarg), maximum (AIRargMAX) and the acute to maximum ratio (acute:MAX) (**Figure 2**). As expected, AIRarg and AIRargMAX were both reduced, suggesting loss of functional and not-fully functional BCM. The ratio acute:MAX is higher in T2D compared to HOV, suggesting that despite loss of both functional and not-fully functional beta cells, a higher proportion of beta cells that are lost those that require maximal stimulation and could possibly be categorized as not-fully functional, stressed or dormant (**Figure 2**).

In the whole pancreas, we found that a model with SUVR-1, as the only imaging metric, in combination with clinical biomarkers, was predictive of acute beta cell function (AIRarg).

SUVR-1, centroid and principal axis length together with clinical biomarkers were predictive of maximum beta cell function (AIRargMAX) in the whole pancreas. This suggests that, at least for T2D, the addition of MRI-based morphology metrics with SUVR-1, improves prediction of structural and functional changes associated with loss of both functional and not-fully functional beta cells for the whole pancreas, compared to PET-only metrics (SUVR-1). Previous histological findings demonstrated that T2D pancreata have greater rates of intralobular fibrosis and acinar to ductal metaplasia than non-diabetic pancreata[35]. Therefore, unlike T1D, where drastic acinar cell volume loss occurs, acinar cells in T2D appear to remodel the pancreas through acinar to ductal metaplasia and increasing fibrosis, in agreement with less severe pancreas volume loss. The inclusion of all three centroid directions and principal axis length 3 suggests that acinar remodeling and fibrosis across the whole pancreas shifts the pancreas center of mass but also shrinks the pancreas to some extent along a short axis in T2D compared to HOV. Previous MRI-metrics in T1D have noted that acinar atrophy typically occurs along the short axes but the long axis remains mostly fixed due to the main duct running the length of the pancreas[29]. Similarly, in our results only principal axis length 3 was the only axis in the whole pancreas that was predictive of AIRargMAX.

In our study, surface area was not predictive of AIRarg or AIRargMAX in our regression models and did not demonstrate group differences between HOV and T2D in the whole pancreas; however, surface area of the pancreas body subregion was significantly lower in T2D (**Figure 7F**). Baseline pancreas volume and pancreas fractal dimension (similar to surface area) were significantly lower in T2D compared to non-diabetic controls[2,4] and at two-year follow-up, those with T2D remission had decreased pancreas fractal dimension and higher pancreas volume[4]. A separate histological evaluation of the pancreas in T2D revealed that a majority of endocrine cell loss occurred in the head and tail with no significant changes in the body[36]. In our dataset, MRI morphology metrics: principal axis length 3, EquivDiameter, bounding box 1, convex volume and surface area demonstrated group differences between HOV and T2D, but only in the pancreas body (**Figure 7**). Suggesting that while limited endocrine loss may be occurring in the pancreas body[36], significant exocrine remodeling in the body may be lead to changes visualized by such MRI morphology metrics. MRI of the pancreas has also been used in T2D to assess anterior-to-posterior diameter on axial slices, similar to our principal axis length 2 or 3 metrics. This method revealed significantly lower anterior-to-posterior pancreas diameters only for body and tail in short term T2D, while long term T2D had lower diameter in all regions (head, body, tail)[31]. Together, suggesting that exocrine changes in the pancreas may occur earlier and more severely in the body of the pancreas, although this remains to be studied longitudinally both at onset and during treatment.

We performed exploratory correlations between single imaging metrics and either AIRarg or AIRargMAX. Pancreas head SUVR-1 and Pancreas body principal axis length 3 were both significantly correlated to AIRarg and AIRargMAX (**Figure 8**). Typically the highest proportion of beta cells are lost from the head in T2D[36] and this was reflected in our previous report where the pancreas head SUVR-1 showed the largest differences between T2D and HOV (-17%)[3]; however, principal axis length 3 in the pancreas body reflecting exocrine cell remodeling and loss in the pancreas body may also be predictive of endocrine cell loss (**Figure 8**).

Several studies have already shown utility of pancreas MRI-based morphology metrics longitudinally in T1D and with the ability to predict outcomes[30,37]. Our current study was performed retrospectively in a cross-sectional cohort of HOV, prediabetes and T2D, and it remains to be seen whether similar patterns and utility occur prospectively in both T2D and T1D combining PET and MRI metrics.

VMAT2 and proinsulin have been shown to be co-expressed and increased amount of VMAT2/proinsulin expression was indicative of larger but dormant beta cells[38], suggesting that VMAT2 may more accurately reflect an insulin vesicle functional capacity reservoir in non-functional and functional beta cells and possibly hybrid alpha-beta-like cells[39]. This might explain the ability of [^18^F]FP-(+)-DTBZ to capture functional and not-fully functional beta cell mass in this cohorts.

To our knowledge, this study is the first to combine PET imaging of BCM and MRI morphology metrics with a robust machine learning-based variable selection method to extract useful PET- and MRI-based metrics for predicting functional and not-fully functional BCM. However, there are several limitations. Given the retrospective nature of the study, it is not possible to determine the temporal sequence of PET and MRI morphology metrics during progression to T2D. Thus, prospective longitudinal studies are necessary. This retrospective study was a relatively small samples size, although typical for PET imaging cohorts. The findings here need to be validated in larger, more diverse etiologies of T2D progression and treatment. Future investigations could also incorporate additional MR imaging to study MR relaxometry, quantitative fat fraction maps, diffusion-weighted imaging, perfusion imaging, MR elastography, for example[27,28], which would allow for further understanding of how changes in pancreas tissue composition drive the morphological changes we observed and how they relate to BCM assessed with PET imaging.

## Conclusion

Applying a robust machine learning-based variable selection method with a multi-modal imaging paradigm, integrating PET with morphological metrics from MRI, provides a more complete understanding of functional and not-fully functional BCM alterations in T2D. This synergistic approach offers a novel combination of biomarkers for staging of pancreatic diseases, such as T2D, and possible methods to evaluate therapeutic interventions.

## Data Availability

All data produced in the present study are available upon reasonable request to the authors.

## Acknowledgments

The authors received support from the National Institutes of Health/National Institute of Diabetes and Digestive and Kidney Diseases (K01DK118005 [JB]) during the writing of this manuscript. The original data acquisition was performed as part of the Pfizer Yale Bioimaging Alliance. The study sponsor/funder was not involved in the writing of the manuscript and did not impose any restrictions regarding the publication of the report.

## Supplementary Data

### Supplementary Table 1

**Supplementary Table 1.**
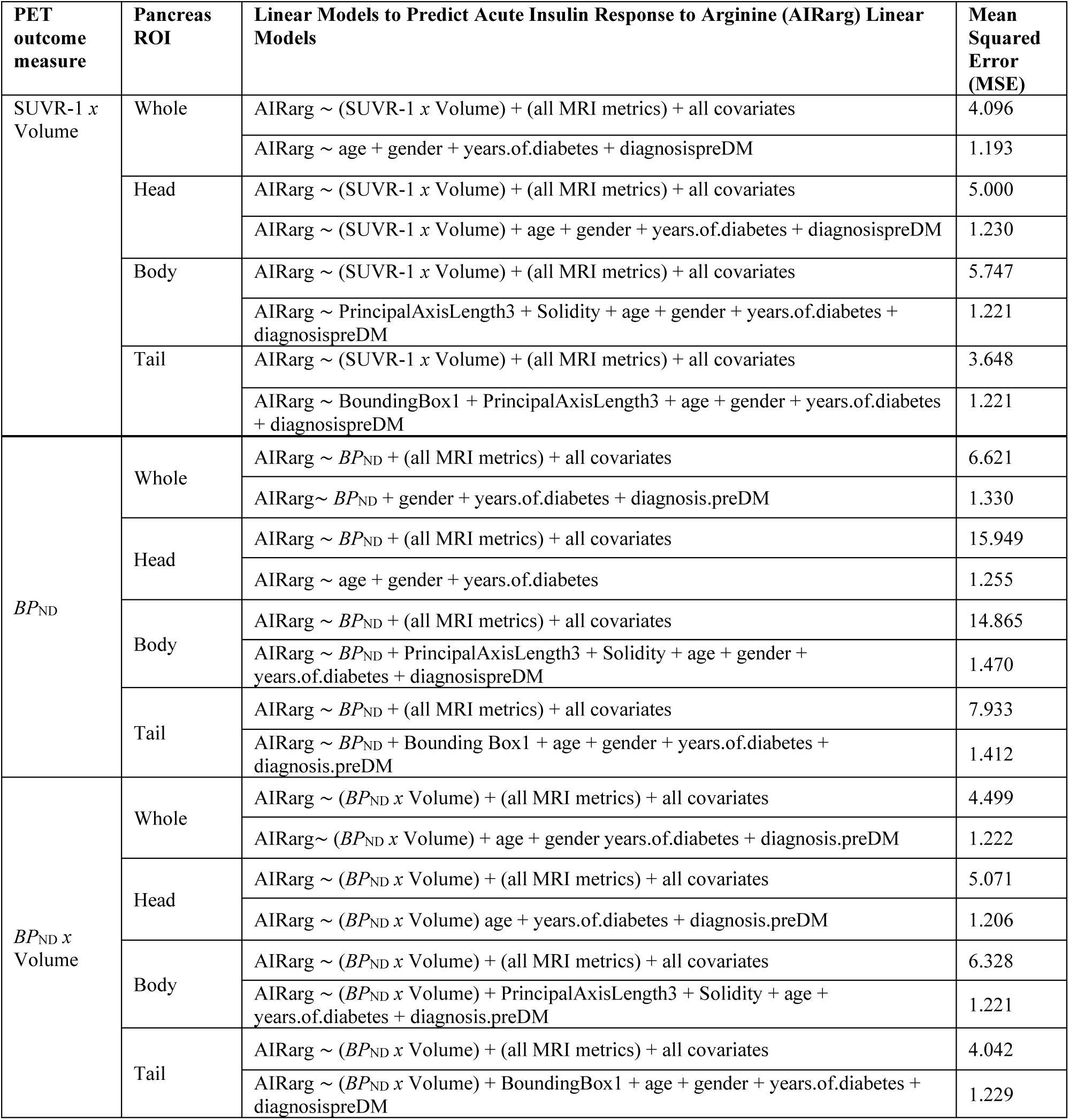

| PET outcome measure | Pancreas ROI | Linear Models to Predict Acute Insulin Response to Arginine (AIRarg) Linear Models | Mean Squared Error (MSE) |
| --- | --- | --- | --- |
| SUVR-1 × Volume | Whole | AIRarg ∼ (SUVR-1 × Volume) + (all MRI metrics) + all covariates | 4.096 |
| | | AIRarg ∼ age + gender + years.of.diabetes + diagnosispreDM | 1.193 |
| | Head | AIRarg ∼ (SUVR-1 × Volume) + (all MRI metrics) + all covariates | 5.000 |
| | | AIRarg ∼ (SUVR-1 × Volume) + age + gender + years.of.diabetes + diagnosispreDM | 1.230 |
| | Body | AIRarg ∼ (SUVR-1 × Volume) + (all MRI metrics) + all covariates | 5.747 |
| | | AIRarg ∼ PrincipalAxisLength3 + Solidity + age + gender + years.of.diabetes + diagnosispreDM | 1.221 |
| | Tail | AIRarg ∼ (SUVR-1 × Volume) + (all MRI metrics) + all covariates | 3.648 |
| | | AIRarg ∼ BoundingBox1 + PrincipalAxisLength3 + age + gender + years.of.diabetes + diagnosispreDM | 1.221 |
| $BP_{ND}$ | Whole | AIRarg $\sim BP_{ND}$ + (all MRI metrics) + all covariates | 6.621 |
| | | AIRarg $\sim BP_{ND}$ + gender + years.of.diabetes + diagnosis.preDM | 1.330 |
| | Head | AIRarg $\sim BP_{ND}$ + (all MRI metrics) + all covariates | 15.949 |
| | | AIRarg ∼ age + gender + years.of.diabetes | 1.255 |
| | Body | AIRarg $\sim BP_{ND}$ + (all MRI metrics) + all covariates | 14.865 |
| | | AIRarg $\sim BP_{ND}$ + PrincipalAxisLength3 + Solidity + age + gender + years.of.diabetes + diagnosispreDM | 1.470 |
| | Tail | AIRarg $\sim BP_{ND}$ + (all MRI metrics) + all covariates | 7.933 |
| | | AIRarg $\sim BP_{ND}$ + Bounding Box1 + age + gender + years.of.diabetes + diagnosis.preDM | 1.412 |
| $BP_{ND} \times$ Volume | Whole | AIRarg $\sim (BP_{ND} \times$ Volume) + (all MRI metrics) + all covariates | 4.499 |
| | | AIRarg $\sim (BP_{ND} \times$ Volume) + age + gender years.of.diabetes + diagnosis.preDM | 1.222 |
| | Head | AIRarg $\sim (BP_{ND} \times$ Volume) + (all MRI metrics) + all covariates | 5.071 |
| | | AIRarg $\sim (BP_{ND} \times$ Volume) age + years.of.diabetes + diagnosis.preDM | 1.206 |
| | Body | AIRarg $\sim (BP_{ND} \times$ Volume) + (all MRI metrics) + all covariates | 6.328 |
| | | AIRarg $\sim (BP_{ND} \times$ Volume) + PrincipalAxisLength3 + Solidity + age + years.of.diabetes + diagnosis.preDM | 1.221 |
| | Tail | AIRarg $\sim (BP_{ND} \times$ Volume) + (all MRI metrics) + all covariates | 4.042 |
| | | AIRarg $\sim (BP_{ND} \times$ Volume) + BoundingBox1 + age + gender + years.of.diabetes + diagnosispreDM | 1.229 |

**Supplementary Table 2.**

| PET outcome measure | Pancreas ROI | Linear Models to Predict maximum insulin response to arginine (AIRargMAX) | Mean Squared Error (MSE) |
| --- | --- | --- | --- |
| SUVr-1 x Volume | Whole | AIRargMAX ~ (SUVr-1 x Volume) + (all MRI metrics) + all covariates | 18.863 |
|  |  | AIRargMAX ~ (SUVr-1 x Volume) + Centroid2 + Centroid3 + PrincipalAxisLength3 + age + gender + BMI + HbA1c + years.of.diabetes + diagnosisT2D | 8.024 |
|  | Head | AIRargMAX ~ (SUVr-1 x Volume) + (all MRI metrics) + all covariates | 18.670 |
|  |  | AIRargMAX ~ (SUVr-1 x Volume) + Centroid1 + Centroid2 + Centroid3 + PrincipalAxisLength2 + PrincipalAxisLength3 + age + gender + HbA1c + years.of.diabetes + diagnosisT2D | 8.220 |
|  | Body | AIRargMAX ~ (SUVr-1 x Volume) + (all MRI metrics) + all covariates | 24.756 |
|  |  | AIRargMAX ~ (SUVr-1 x Volume) + Centroid2 + Centroid3 + PrincipalAxisLength3 + age + gender + HbA1c + years.of.diabetes + diagnosisT2D | 6.474 |
|  | Tail | AIRargMAX ~ (SUVr-1 x Volume) + (all MRI metrics) + all covariates | 26.988 |
|  |  | AIRargMAX ~ (SUVr-1 x Volume) + Centroid1 + Centroid3 + age + gender + HbA1c + years.of.diabetes + diagnosisT2D | 7.524 |
| BP<sub>ND</sub> | Whole | AIRargMAX ~ BP<sub>ND</sub> + (all MRI metrics) + all covariates | 44.235 |
|  |  | AIRargMAX ~ BP<sub>ND</sub> + Centroid2 + Centroid3 + PrincipalAxisLength3 + gender + HbA1c + years.of.diabetes + diagnosisT2D | 7.743 |
|  | Head | AIRargMAX ~ BP<sub>ND</sub> + (all MRI metrics) + all covariates | 27.363 |
|  |  | AIRargMAX ~ BP<sub>ND</sub> + Centroid2 + Centroid3 + PrincipalAxisLength2 + PrincipalAxisLength3 + gender + HbA1c + years.of.diabetes + diagnosisT2D | 8.271 |
|  | Body | AIRargMAX ~ BP<sub>ND</sub> + (all MRI metrics) + all covariates | 83.606 |
|  |  | AIRargMAX ~ BP<sub>ND</sub> + Centroid1 + Centroid2 + Centroid3 + PrincipalAxisLength3 + age + gender + HbA1c + years.of.diabetes + diagnosisT2D + diagnosispreDM | 7.325 |
|  | Tail | AIRargMAX ~ BP<sub>ND</sub> + (all MRI metrics) + all covariates | 82.360 |
|  |  | AIRargMAX ~ BP<sub>ND</sub> + Centroid1 + Centroid3 + PrincipalAxisLength3 + HbA1c + years.of.diabetes + diagnosisT2D | 7.688 |
| BP<sub>ND</sub> x Volume | Whole | AIRargMAX ~ (BP<sub>ND</sub> x Volume) + (all MRI metrics) + all covariates | 18.141 |
|  |  | AIRargMAX ~ (BP<sub>ND</sub> x Volume) + Centroid2 + Centroid3 + PrincipalAxisLength3 + age + gender + BMI + HbA1c + years.of.diabetes + diagnosisT2D | 8.167 |
|  | Head | AIRargMAX ~ (BP<sub>ND</sub> x Volume) + (all MRI metrics) + all covariates | 15.556 |
|  |  | AIRargMAX ~ (BP<sub>ND</sub> x Volume) + Centroid2 + Centroid3 + PrincipalAxisLength2 + PrincipalAxisLength3 + age + gender + HbA1c + years.of.diabetes + diagnosisT2D | 8.071 |
|  | Body | AIRargMAX ~ (BP<sub>ND</sub> x Volume) + (all MRI metrics) + all covariates | 23.425 |
|  |  | AIRargMAX ~ (BP<sub>ND</sub> x Volume) + Centroid2 + Centroid3 + PrincipalAxisLength3 + age + gender + HbA1c + years.of.diabetes + diagnosisT2D | 6.469 |
|  | Tail | AIRargMAX ~ (BP<sub>ND</sub> x Volume) + (all MRI metrics) + all covariates | 28.240 |
|  |  | AIRargMAX ~ (BP<sub>ND</sub> x Volume) + Centroid1 + Centroid3 + age + gender + HbA1c + years.of.diabetes + diagnosisT2D | 7.169 |

**Supplementary Table 3.**

| PET outcome measure | Pancreas ROI | Linear Models to Predict ratio of acute to maximum insulin response to arginine (acute:MAX) | Mean Squared Error (MSE) |
| --- | --- | --- | --- |
| SUV<sub>R</sub>-1 x Volume | Whole | acute:MAX ~ (SUV<sub>R</sub>-1 x Volume) + (all MRI metrics) + all covariates | 0.053 |
|  |  | acute:MAX ~ Centroid1 + Centroid3 + BoundingBox1 + BoundingBox3 + PrincipalAxisLength1 + PrincipalAxisLength3 + age + gender + HbA<sub>1c</sub> + years.of.diabetes + diagnosisT2D + diagnosispreDM | 0.023 |
|  | Head | acute:MAX ~ (SUV<sub>R</sub>-1 x Volume) + (all MRI metrics) + all covariates | 0.096 |
|  |  | acute:MAX ~ PrincipalAxisLength1 + PrincipalAxisLength2 + PrincipalAxisLength3 + age + HbA<sub>1c</sub> + years.of.diabetes + diagnosisT2D + diagnosispreDM | 0.024 |
|  | Body | acute:MAX ~ (SUV<sub>R</sub>-1 x Volume) + (all MRI metrics) + all covariates | 0.034 |
|  |  | acute:MAX ~ Centroid1 + Centroid2 + Centroid3 + BoundingBox1 + BoundingBox3 + Extent + PrincipalAxisLength2 + Solidity + age + gender + weight + HbA<sub>1c</sub> + years.of.diabetes + diagnosisT2D + diagnosispreDM | 0.022 |
|  | Tail | acute:MAX ~ (SUV<sub>R</sub>-1 x Volume) + (all MRI metrics) + all covariates | 0.072 |
|  |  | acute:MAX ~ Centroid1 + Centroid2 + PrincipalAxisLength2 + age + HbA<sub>1c</sub> + years.of.diabetes + diagnosisT2D + diagnosispreDM | 0.025 |
| BP<sub>ND</sub> | Whole | acute:MAX ~ BP<sub>ND</sub> + (all MRI metrics) + all covariates | 0.105 |
|  |  | acute:MAX ~ Centroid1 + BoundingBox1 + BoundingBox3 + PrincipalAxisLength3 + gender + HbA<sub>1c</sub> + years.of.diabetes + diagnosispreDM | 0.023 |
|  | Head | acute:MAX ~ BP<sub>ND</sub> + (all MRI metrics) + all covariates | 0.343 |
|  |  | acute:MAX ~ PrincipalAxisLength2 + PrincipalAxisLength3 + gender + HbA<sub>1c</sub> + years.of.diabetes | 0.022 |
|  | Body | acute:MAX ~ BP<sub>ND</sub> + (all MRI metrics) + all covariates | 0.074 |
|  |  | acute:MAX ~ BP<sub>ND</sub> + Centroid1 + Centroid2 + Centroid3 + BoundingBox2 + BoundingBox3 + Extent + PrincipalAxisLength2 + Solidity + age + gender + weight + BMI + HbA<sub>1c</sub> + years.of.diabetes + diagnosispreDM | 0.022 |
|  | Tail | acute:MAX ~ BP<sub>ND</sub> + (all MRI metrics) + all covariates | 0.186 |
|  |  | acute:MAX ~ Centroid1 + HbA<sub>1c</sub> + years.of.diabetes | 0.025 |
| BP<sub>ND</sub> x Volume | Whole | acute:MAX ~ (BP<sub>ND</sub> x Volume) + (all MRI metrics) + all covariates | 0.055 |
|  |  | acute:MAX ~ Centroid1 + BoundingBox1 + BoundingBox3 + PrincipalAxisLength1 + PrincipalAxisLength3 + age + HbA<sub>1c</sub> + years.of.diabetes + diagnosisT2D + diagnosispreDM | 0.020 |
|  | Head | acute:MAX ~ (BP<sub>ND</sub> x Volume) + (all MRI metrics) + all covariates | 0.105 |
|  |  | acute:MAX ~ PrincipalAxisLength1 + PrincipalAxisLength2 + PrincipalAxisLength3 + age + HbA<sub>1c</sub> + years.of.diabetes + diagnosisT2D + diagnosispreDM | 0.024 |
|  | Body | acute:MAX ~ (BP<sub>ND</sub> x Volume) + (all MRI metrics) + all covariates | 0.036 |
|  |  | acute:MAX ~ (BP<sub>ND</sub> x Volume) + Centroid1 + Centroid2 + Centroid3 + BoundingBox1 + BoundingBox3 + Extent + PrincipalAxisLength2 + Solidity + age + gender + HbA<sub>1c</sub> + years.of.diabetes + diagnosisT2D + diagnosispreDM | 0.023 |
|  | Tail | acute:MAX ~ (BP<sub>ND</sub> x Volume) + (all MRI metrics) + all covariates | 0.067 |
|  |  | acute:MAX ~ Centroid1 + Centroid2 + PrincipalAxisLength2 + age + HbA<sub>1c</sub> + years.of.diabetes + diagnosisT2D + diagnosispreDM | 0.025 |

**Supplementary Figure 1.**
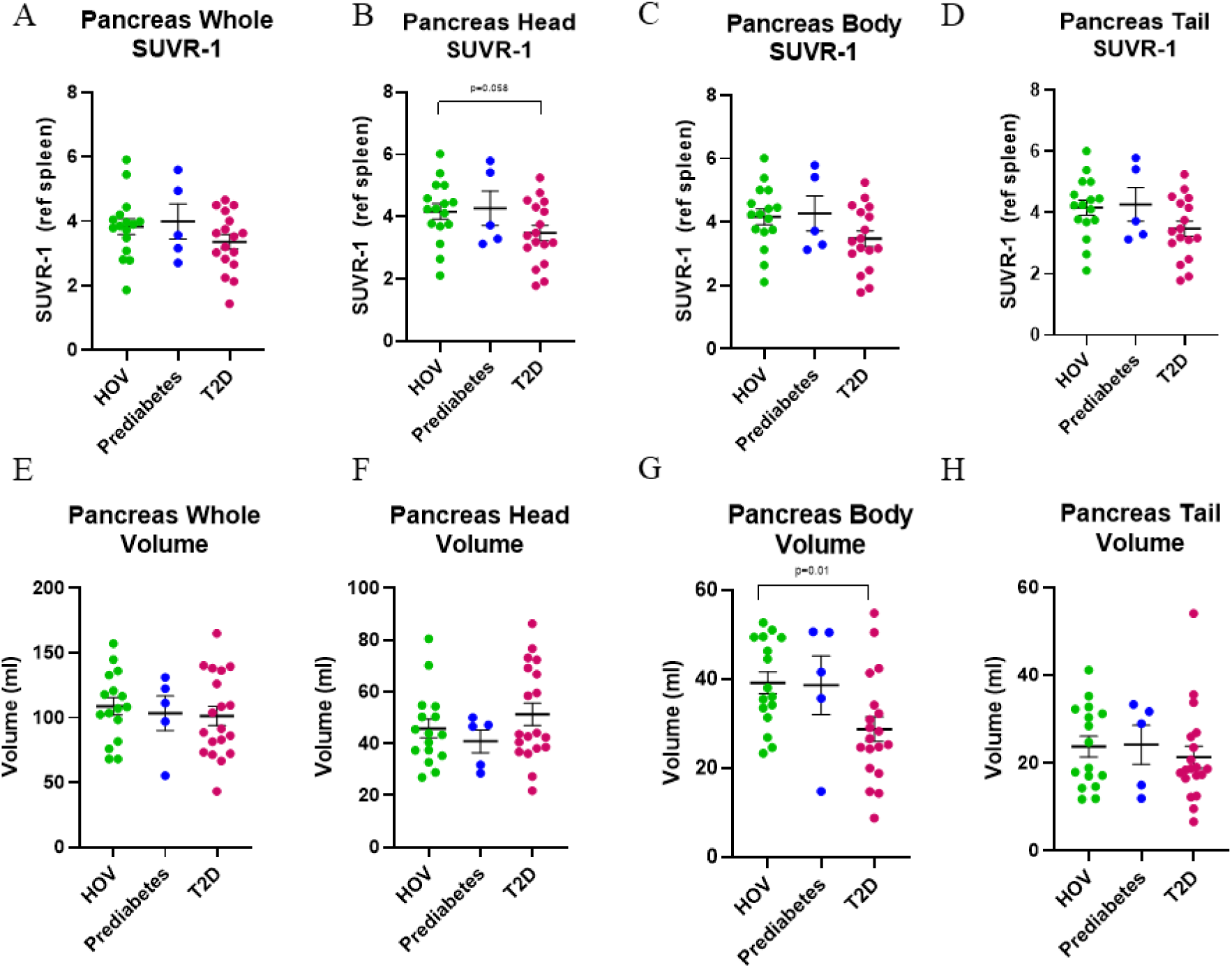
Group comparisons between healthy obese volunteers and individuals with T2D of the PET outcome metric SUVR-1 in pancreas **A)** whole **B)** head **C)** body and **D)** tail. Pancreas MRI volume metrics for pancreas **E)** whole **F)** head **G)** body and **H)** tail. All data presented as mean ± SEM.

